# Forecasting Covid-19 Outbreak Progression in Italian Regions: A model based on neural network training from Chinese data

**DOI:** 10.1101/2020.04.09.20059055

**Authors:** Cosimo Distante, Igor Gadelha Pereira, Luiz M. Garcia Gonçalves, Prisco Piscitelli, Alessandro Miani

## Abstract

**Background:** Epidemiological figures of Covid-19 epidemic in Italy are worse than those observed in China.

**Methods:** We modeled the Covid-19 outbreak in Italian Regions vs. Lombardy to assess the epidemics progression and predict peaks of new daily infections and total cases by learning from the entire Chinese epidemiological dynamics. We trained an artificial neural network model, a modified auto-encoder with Covid-19 Chinese data, to forecast epidemic curve of the different Italian regions, and use the susceptible–exposed–infected–removed (SEIR) compartment model to predict the spreading and peaks. We have estimated the basic reproduction number (R_0_) - which represents the average number of people that can be infected by a person who has already acquired the infection - both by fitting the exponential growth rate of the infection across a 1-month period, and also by using a day by day assessment, based on single observations.

**Results:** The expected peak of SEIR model for new daily cases was at the end of March at national level. The peak of overall positive cases is expected by April 11^th^ in Southern Italian Regions, a couple of days after that of Lombardy and Northern regions. According to our model, total confirmed cases in all Italy regions could reach 160,000 cases by April 30^th^ and stabilize at a plateau.

**Conclusions:** Training neural networks on Chinese data and use the knowledge to forecast Italian spreading of Covid-19 has resulted in a good fit, measured with the mean average precision between official Italian data and the forecast.

## Introduction

According to the Italian National Institute of Health (ISS), at the date of April 8^th^ in Italy there were about 140,000 people positive to the 2019-nCoV (including deceased patients) since the beginning of epidemic (95,262 currently positive and 26,491 healed).[7] About 53% of cases are males (median age: 62 years old). Detailed epidemiological figures are provided by the Italian National Institute of Health (ISS) and tell us that men represent the majority of cases in people aged 0-9 and 50-79 (range 52-63%), while in the younger age groups 0-19, as well as between 80 and 89 years old, males and females are equally represented among people who tested positive for Covid-19. Women accounted for 70% of cases >90 years old and about 55% between 20 and 39 years of age, but men represented also the vast majority of deceased people in all the age groups up to 89 years old (range 57-79%).[7]

Regional figures are available up to April 8^th^ and show that about 30% (n=28,545) of currently positive people still live in Lombardy (56% if considering the overall cases confirmed from the beginning of the epidemic), followed by Emilia Romagna (13.7% of currently positive people), Piedmont (11.5%), Veneto (n=10.7%), Tuscany (5.8%), Marche (3.7%), Lazio (n=3.6%), Liguria and Trentino Alto Adige (3.4%), Campania and Apulia (3%), Sicily (2%), Friuli Venezia Giulia and Abruzzo (1.5%), and less than 1% in Umbria, Sardinia, Calabria, Val d’Aosta, Basilicata, and Molise. [7]

A total of 28,485 symptomatic people were hospitalized at the same date of April 8^th^ in Italy, with Lombardy accounting for 11,719 hospital admissions (41%), followed by Emilia Romagna (n=3,769), Piedmont (n=3,493) and Veneto (n=1,554). Only Tuscany (n=1,066), Liguria (n=1,109) and Lazio (n=1,241) Marche (n=974), recorded more than 600 hospital admissions at regional level, while other regions remain still lower. At the present, 3,693 patients are assisted in intensive care units (34% only in Lombardy), with a reduction of about 8% in the first week of April. A total of 1,257 ICU patients were in Lombardy (11% of hospitalized people), 423 in Piedmont (12%), 285 in Veneto (18%), 361 in Emilia Romagna (9%), 260 in Tuscany, 163 in Liguria, 140 in Trentino Alto Adige, and 133 in Marche region. With the exception of Lazio (n=196), Campania (n=97) and Apulia (n=90), all the other regions of Central and Southern Italy, at the moment have less than 65 patients admitted to the ICUs of their regional healthcare systems. [7]

On April 8^th^, total deaths were 17,669 at national level (+65% from March 30^th^ to April 8^th^), with 9,722 in Lombardy (56%), 2,234 in Emilia Romagna (13%), 1,378 in Piedmont (8%), 736 in Veneto (4%), 652 in Marche region (3.6%), 654 in Liguria, 438 in Trentino Alto Adige, 392 in Tuscany, 244 in Lazio, 221 in Campania, 219 in Apulia, 179 in Abruzzo, 169 in Friuli Venezia Giulia, 133 in Sicily, 102 in Val d’Aosta and less than 50 in the other four regions (Figure 1).[7] Lethality rates seems to increase with age and it is higher in males: 0% from 0 to 29 and <1% between 30 and 49 years of age; 2.3% in the age group 50-59 (1% in women and 3.5% in men); 8.4% from 60 to 69 years old (5.1% in women and 10.3% in men); 22.7% from 70 to 79 (15.6% in females and 26.9% in males); 30.6% from 80 to 89 years old (22.6% and 38.4% in men); 26.8% after 90 years old (21.2%in females and 40.6% in males).[7] In Fig.1 we report the PCR testing rate and mortality per region.

**Figure 1.**
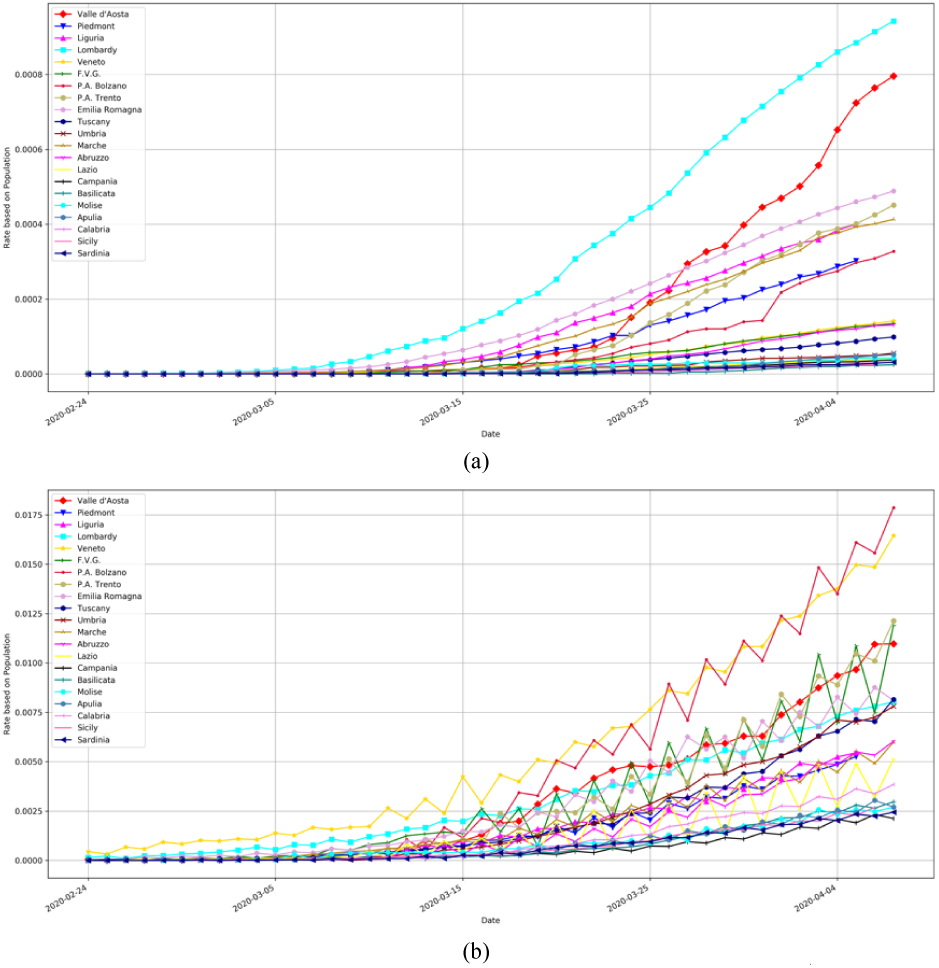
Mortality (a) and PCR test (b) rate in Italy per region at the date of April 8^th^.

Based on these figures, it is clear that the Covid-19 outbreak is now putting overwhelming pressure mainly on Lombardy and Northern regions of the Po Valley (Padana plain), but the peak of the epidemics has not yet been reached. Until now, Southern Regions seem to be less affected by the Covid-19 infection although a huge number of people - mainly students attending Universities in Northern Italy - came back from Po Valley to their families in the South just in the middle of the outbreak, thus representing a potential factor able to accelerate the spreading of the viral infection. The peaks of the epidemics for new daily cases have been reached in the majority of regions, but peaks of total positive cases is expected to be delayed in Central and Southern Italian regions compared to Lombardy and Northern ones. Here we present an attempt to predict the peak of the outbreak in Italy, which was confirmed at national level at the end of March in terms of new daily cases, with an expected subsequent different progression of the epidemics in Southern Italian Regions compared to Lombardy in terms of total cases.

### Methods: Modeling the Covid-19 outbreak progression in Italian Regions

The correct prediction of new daily cases at this time of Italian COVID-19 outbreak requires the correct estimation of the peak including the unknown remaining part of the epidemiological curve, where this later can be predicted ahead by using a deep convolutional auto-encoder. Therefore, we applied a Modified Auto-Encoder (MAE) for a time-series forecast in order to predict the evolution of daily cases for each of the 21 regions of Italy. [15] The model was trained with the data from the Chinese regions, which provides complete data in the sense that they already went through the peak number of daily cases and managed to suppress the epidemic by social distancing measures. Although such measures implemented by the Chinese government may be impossible to implement in other countries or may not be as effective as it was in China, the data generated by their experience going through the epidemic can be used to derive data-oriented models to predict the epidemic dynamic behavior in other countries. The forecast of new daily cases has been used to correctly estimate the single peaks but also to obtain better spreading predictions from SEIR model. We modeled spreading of Covid-19 using Chinese data and used the model to predict epidemic curve in each Italian region, allowing to gain better information on the new daily cases peaks with the forecasted curve. The forecast portion of the curve allows to have a better prediction of active cases with the SEIR model, by computing the position of the peaks of active cases for each Italian region. We present the data sources used to train the model, the architecture of the model and the details about the training and forecasting procedures. In the end, we present the results obtained for the forecasting of daily cases for Lombardy and the biggest Southern Italian regions, as well as for all other regions.

### Data Source

We used the dataset provided by the Johns Hopkins University Center for Systems Science and Engineering (JHU CSSE) that is sourced from the World Health Organization (WHO). The dataset provides the number of confirmed cases for each country since January 22, the day that the WHO was notified of an epidemy caused by a new Coronavirus in China.

From January 22^nd^up to February 13^th^, the Chinese government reports only the cases that were confirmed by laboratory tests as confirmed cases of the Covid-19 disease. After February 14, the Chinese government starts to consider not only the laboratory-confirmed cases but also the cases diagnosed by clinical procedures. On February 14^th^, in the Hubei province, the epicenter of the Coronavirus epidemic in China, the number of clinically confirmed cases was 15,384 and the number of laboratory-confirmed cases was 36,602. Hence, the total number of confirmed cases from January 22 up to February 13 was adjusted by the Equation

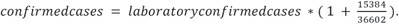

The data provided by the dataset regards the total number of cases confirmed up to date. However, to use the daily cases, we differentiate the data of each region with regards to each day.

In order to train our model, we used the data from 31 Provinces/Cities of mainland China and the three other regions: Hong Kong, Macau, and Taiwan. In total, data from 34 regions were used and represented on the columns of the matrix *D* with rows representing each day since the first case reported on the region.

The Italian regional time series used in this paper for the forecast, has been taken from the Italian Department for Citizens’ Protection [17].

### Modeling Time Series

We used aModified Auto-Encoder (MAE) to forecast the number of confirmed daily cases of Covid-19 disease in Italy regions. Traditional auto-encoders [16]models the input/output data by creating a hidden probabilistic representation of the data in its middle layer, also called latent space. The MAE model modifies the traditional auto-encoders to employ an extra output branch derived from the latent space. While the traditional output of auto-encoder architectures is designed to be trained to match the input, in our case, the extra output is modified to predict also the next sample of the sequencetime series given to the input. Figure 2 depicts the MAE model architecture.

**Figure 2.**
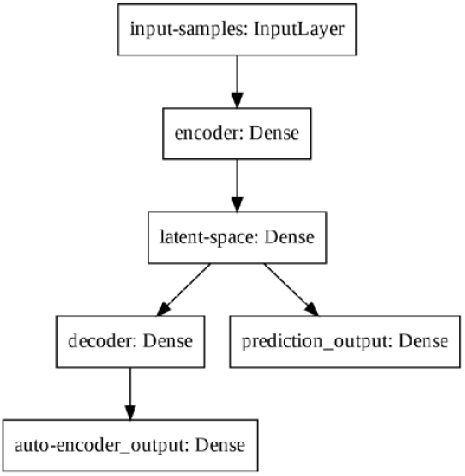
Modified Auto-Encoder Architecture

The prediction output has one unit while the number of units in the input-samples, encoder, latent-space, decoder, and auto-encoder output is 8, 32, 4, 32, 8, respectively. We randomly select a sequential set of samples of 8 days as a training example and combines 34 examples in one training step, one for each region. Each example is normalized by dividing its values by their mean.

### Forecasting *the new daily cases*

A total of five models are trained with the same dataset and the average of each forecasted prediction is taken as the final prediction of new daily cases. Note that each predicted value must be denormalized by multiplying its value by the mean of the input segment.

The forecasting procedure for a known input segment is generally described as one-step forecasting and intends to evaluate how close the predictions are from the expected values. However, it is new daily cases would occur since it indicates the decrease in the number of cases and a possible end of the epidemic.

Therefore, we use the output of the prediction of step *i-th* as an input to the prediction of the (*i+1)-th*. Repeating such a recursive procedure multiple times gives us a multiple-step ahead forecasting. In this way, we divided the forecast procedures into two phases.

The first phase regards the time in which the real data is available, for this phase we compute only the one-step-ahead forecasting. The second phase regards the time in which there is no real data available, hence we compute multiple-step-ahead forecasting. In general, the second phase starts on the last day of available data.

We have chosen to train the modified auto-encoder by using several data and several latent variables: using only China data and latent variables z=4,8,16; or using only Lombardy data, or all nations data. All the model has been evaluated with the mean average precision on forecasting, and the Figure 3 shows the MAPE computed on all Italian regions:

**Figure 3.**
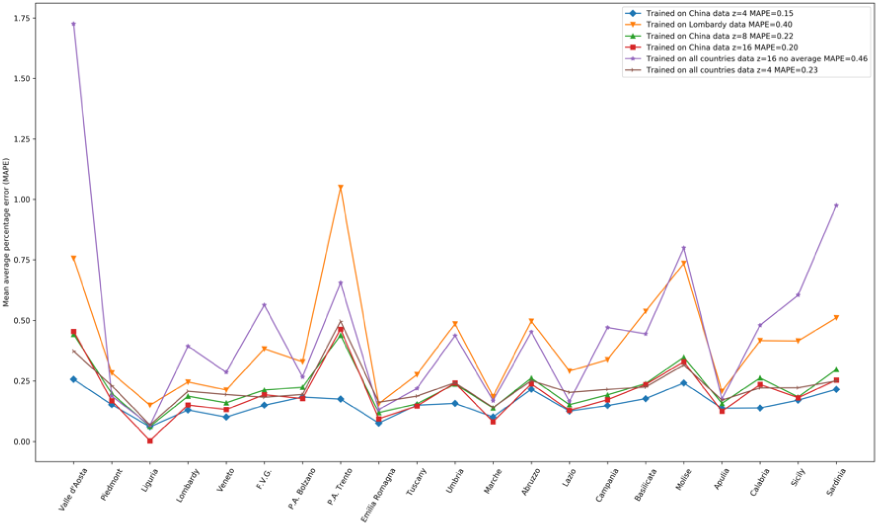
mean average percentage error computed on all regions of the different datasets and latent spaces z=4,8,16 experimented. From the results, data of the provinces of the Chinese data with z=4 provides the optimal model.

The model trained with data from the provinces of China with z=4 presented optimal results. The following results presented at this section are obtained with the optimal model.

We also evaluate the performance of our model by analyzing how close it predicts the next sample in the first phase of the forecasting procedure, that is when real data is available. In the second phase, we evaluate if the model follows a plausible trend for the new daily cases and for the total cumulative cases.

Figure 4a depicts the daily cases and Figure 4b depicts the cumulative cases of Covid-19 for the Lombardy region. From Figure 4a, we can verify that the model is following the real data on the one-step-ahead forecasting and Figure 4b indicates a plausible trend in the multiple-step-ahead forecasting.

**Figure 4.**
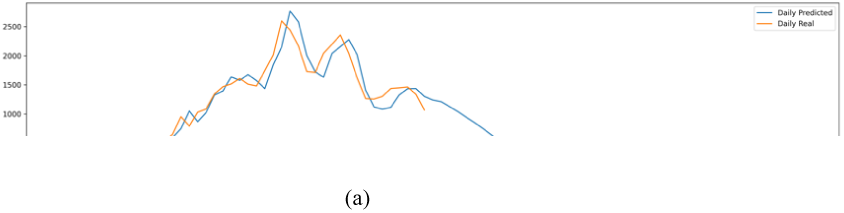
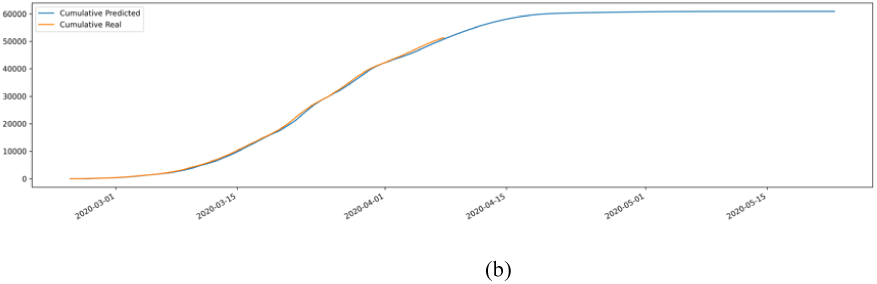
Lombardy prediction of newly daily cases in (a) and the cumulative in (b)

We notice a very similar behavior for the forecasting of daily and cumulative cases for the region of Apulia, depicted by Figure 5a and Figure 5b. In fact, the forecasted daily cases for the region of Apulia decrease much faster than on Lombardy, which makes sense given that Apulia does not accumulate a very high number of cases as Lombardy doesand the social distancing measures were implemented in the early days of the infection.

**Figure 5.**
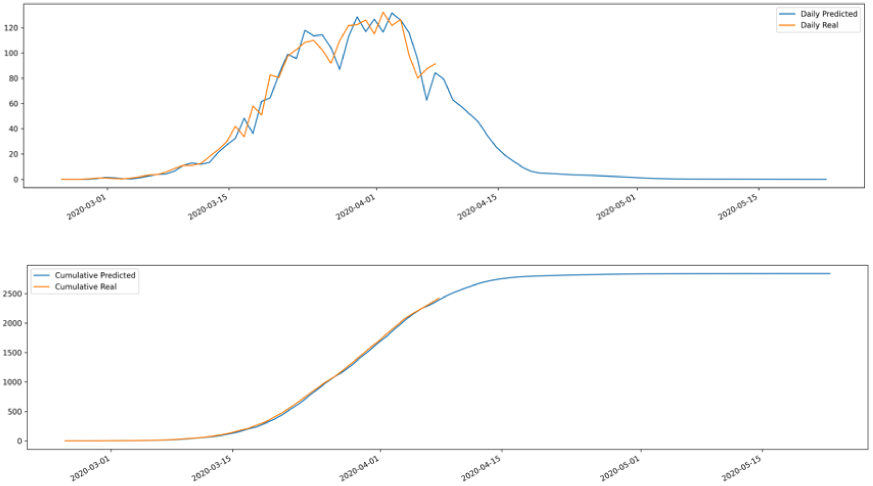
Apulia prediction of newly daily cases in (a) and cumulative in (b)

The last set of Figures, Figure 6a and Figure 6b depict the daily and cumulative cases for all regions of Italy. The idea here is to evaluate how the forecasted values for each region would make sense in a broader scenario. Indeed, the predicted values are very close to the real values. The trend also indicates that the sum of confirmed cases from all Italy regions could reach 160,000 cases by the end of April and stabilize at a plateau.

**Figure 6.**
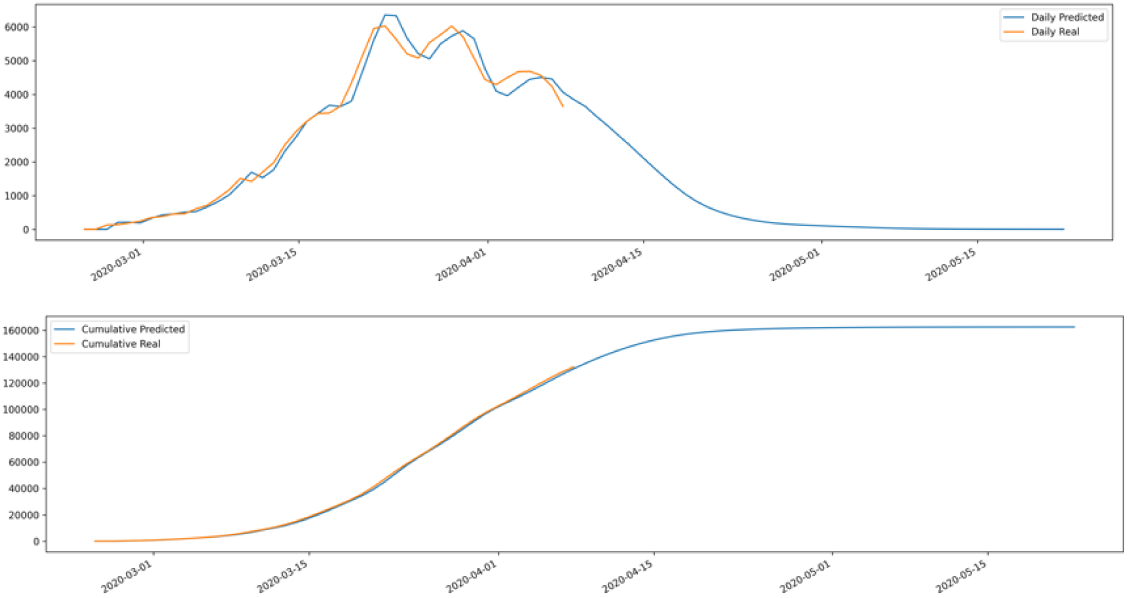
Forecast for Italy data in (a) the new daily cases and in (b) the cumulative distribution

**Figure 7.**
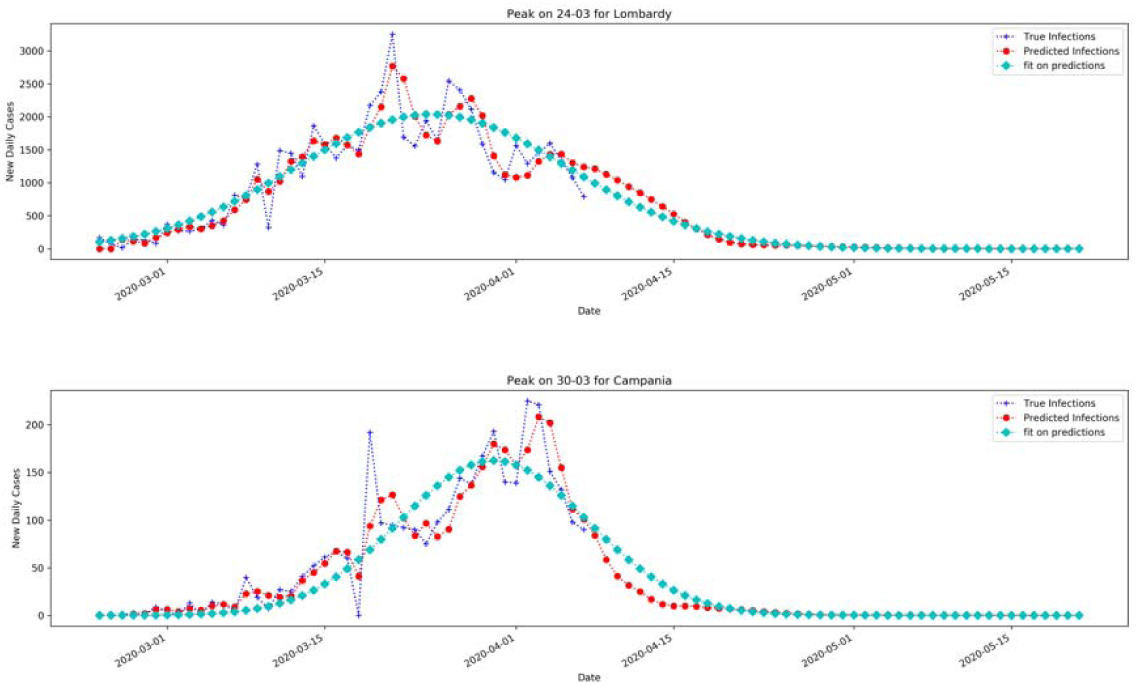
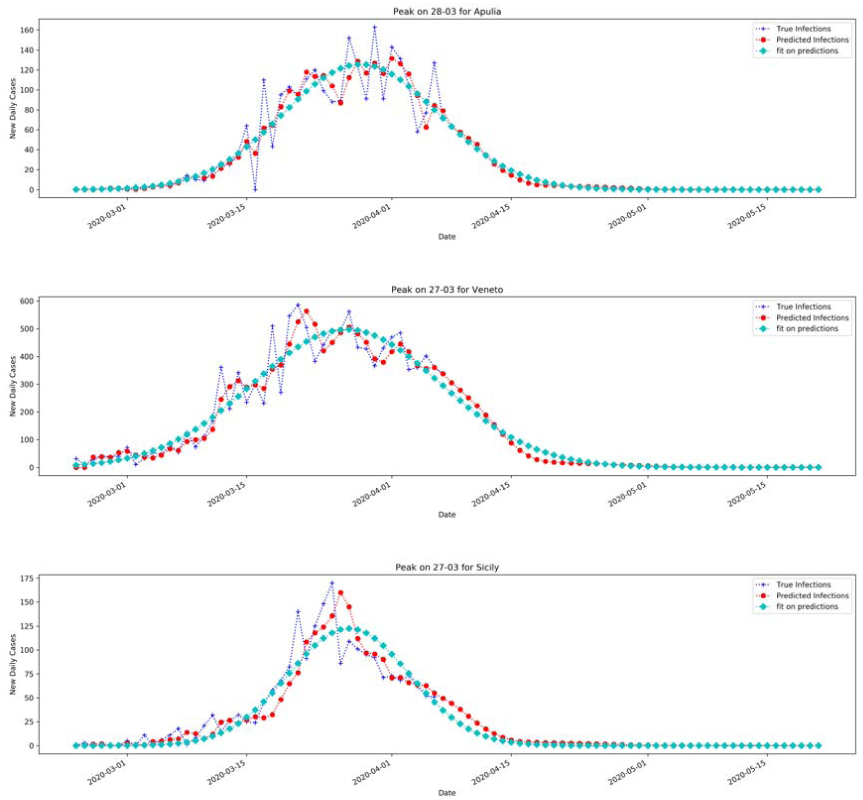
Fit with Gaussian function of the new daily cases with the forecasted samples from April 8^th^ ahead.

With these results we estimated the new daily peak by fitting the forecasted curve with a Gaussian fit to find out the day at which it occurs

### Modeling basic reproductive number R_0_

The basic reproduction number (R0) is an indicator that resumes the average number of people that can be infected by a person who has already acquired the infection. R_0_ is a metric of how contagious is the disease and its correct estimation is extremely important for epidemiologists, especially when facing new diseases like COVID-19. R_0_ can be computed in different ways. In our models, we have estimated the basic reproduction number (R_0_) both by fitting the exponential growth rate of the infection across a 1-month periodand also by using day by day assessment, based on single observations [1]. This study makes use of the susceptible–exposed–infected–removed (SEIR) compartment model [4] to predict the spreading of the pandemic in Italy. Our efforts could be helpfulin the adoption of all the possible preventive measures, and to study of the epidemics progression across Southern regions as opposed to the national trend. This metric can be biased by the optimal estimation of the basic reproductive number R_0_ (pronounced R-nought). It must be said that R_0_ is important if correlated with weather conditions and that reproductive index is reduced as the air temperature and relative humidity increase,[5] according to the formula:

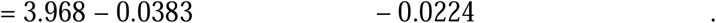

This means the transmission of COVID-19 candecrease with the warmer season, and that at least some

### Daily basis estimation of the reproductive number

R_0_ is an average value, but it can be also computed day by day to monitor the transmission of the infection. Being an average value, it can be skewed by super-spreader events. A super-spreader is an infected individual who infects an unexpectedly large number of people. In Italy this event can be also generated not necessarily by an individual, but from the perturbation of a susceptible population, as it happened in Apulia and Sicily with uncontrolled large group of people coming from outbreak areas. For a “super spreader” individual, such events are not necessarily a bad sign, because they can indicate that fewer people are perpetuating an epidemic. Super-spreaders may also be easier to identify and contain, since their symptoms are likely to be more severe. In short, R_0_ is a moving target. Tracking every case and the transmission of a disease is extremely difficult, so the estimation of R_0_ is a complex and challengingissue: estimates often change as new data becomes available. In [10] a review of 12 studies on the reproduction number for a time period covered from 1 January 2020 to 7 February 2020was analysed for Covid-19 from China and overseas. The work found that the R_0_estimates ranges from1.4 to 6.49[12][11], passing through[14] of 4.08 in mainland China. The review in [10] pointed out a mean of 3.28, a median of 2.79 and interquartile range (IQR) of 1.16, which is considerably higher than the WHO estimate at 1.95. Also in [18]the R_0_for 2019-nCov is reported in the range [1.4, 5.5]. These estimates of R_0_ depend on the estimation method used as well as the validity of the underlying assumptions. In a beginning stage, due to a small amount of data and short time onset, these estimates can be biased, and in a longer period converges to the WHO estimate. The initial estimates result in a reproduction number for Covid-19 higher than SARS coronavirus, where this last it is reported to range between 2 and 5.

If we define the Y(t) as the number of infected people with symptom at time t, the exponential growth rate is *λ* = ln(*Y*(*t*)/*t*).

Let us consider *T*_*g*_ = 7.5 the generation time (i.e. the serial interval) and *T*_*l*_ = 5.2 the latent or incubation time (values taken from [6]). The infectious time *T*_*i*_ = *T*_*g*_ - *T*_*l*_, and the ratio of exposed period to generation time is *ρ*= *T*_*l*_/*T*_*g*_. The basic reproductive number can be approximated to:

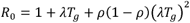

In order to estimate *R*_0_, it is important to find *λ* and then the number of infected people:

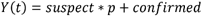

Suspects correspond to the number of individuals screened with test which have been confirmed.

Table 1 shows the *R*_0_estimated values, computed on a daily basis for the Italian regions and for the initial outbreak region Lombardy, where about 40 cases were confirmed out of 100 suspects (Figure 4).The two methods provide agreements of values, although the first method based on exponential fit should provide a better estimation, being computed on the entire time series. From Figure 3, it comes an important aspect with respect to the Wuhan *R*_0_ = 2.68 as reported in [6].

**Table 1.** Estimated R_0_ values of the analyzed regions. Given the trend, min values are usually referred to the beginning of the infection

| Region | $R_0$ | CI 95% | |
| --- | --- | --- | --- |
| Valle d'Aosta | 1.70 | 1,67 | 1,72 |
| Piedmont | 2.15 | 2.11 | 2.18 |
| Lombardy | 3.10 | 3.08 | 3.11 |
| Liguria | 2.04 | 2.02 | 2.06 |
| P.A. Bolzano | 1.45 | 1.41 | 1.50 |
| P.A. Trento | 1.87 | 1.84 | 1.90 |
| Friuli .V.G. | 1.95 | 1.93 | 1.96 |
| Veneto | 2.55 | 2.54 | 2.56 |
| Emilia Romagna | 2.64 | 2.63 | 2.64 |
| Marche | 2.17 | 2.15 | 2.19 |
| Umbria | 1.64 | 1.68 | 1.75 |
| Tuscany | 2.01 | 1.98 | 2.04 |
| Lazio | 1.82 | 1.78 | 1.85 |
| Abruzzo | 1.63 | 1.60 | 1.66 |
| Campania | 1.96 | 1.95 | 1.98 |
| Molise | 1.48 | 1.47 | 1.49 |
| Apulia | 1.73 | 1.70 | 1.76 |
| Basilicata | 1.34 | 1.32 | 1.37 |
| Calabria | 1.47 | 1.43 | 1.50 |
| Sicily | 1.67 | 1.62 | 1.71 |
| Sardinia | 1.64 | 1.61 | 1.67 |

### Modeling transmission in Italy

We used the susceptible-exposed-infectious-recovered (SEIR) model [4] to simulate epidemic since it was established on January 2020. This model is used with the predictions from the modified auto-encoder neural network to better estimate the peaks. It is based on a previous model SIR which was based on three compartments, but since the infection has an incubation period, the compartment E (Exposed) is included. These compartments are modeled over the time, and capture the changes in the population. Let us say that given N the total population, then N=S+E+I+R, where:

- “S” Susceptible is the portion of population that does not have any vax coverage or immune;
- “E” exposed: is the portion of the population that have been infected, but are in the incubation period that do not infect others;
- “I” Infectious: is the portion of N that is infectious and may infect others, they become dead or may recover;
- “R” Recovered: number of infectious people who have been healed, and become immune.

This model captures dynamics of these compartments over the time by four ordinary differential equations. One of the most important aspects of these ordinary differential equations is equilibrium, who is achieved by setting to zero their derivatives along the time t. The two equilibriums are: disease-free equilibrium (DFE) and endemic equilibrium (EE).

Besides equilibrium, stability is an issue who is correlated with the basic reproductive number, where if *R*_0_ <1 DFE is stable; while when *R*_0_ >1 DFE is unstable and EE is stable. The four equations are:

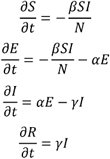

Where *β* = *R*_0_/*T*_*i*_, *α* = 1/*T*_*l*_ and y= 1/*T*_*i*_ with *T*_*i*_ and *T*_*l*_ as defined above, the serial and incubation period respectively. The contact rate *β* is the rate of infection from an infected individual to one of their susceptible contacts on the unitary time step *dt*. The number of individuals transferred from Susceptible state to Exposed state is 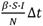. The force of infection is defined as *β* * *S*(*t*)/*N* which is the number of new infections divided the Italian population. At the same time step, there are *αE*(*t*)Δ*t* number of cases that are transferred from Exposed to Infectious compartment, and *γI*(*t*)Δ*t* number of cases transferred from Infectious compartment to Removed. It is important to state that we assume a closed population, which means the population is fixed, then no births, no deaths or introduction of new individuals. From the above ODE system, 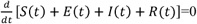 which means that the population N is constant at any time step *t*: S(*t*) + *E*(*t*) + *I*(*t*) + *R*(*t*) = *N* for any *t* ≥ 0.The individuals in exposed state is is infected but not yet infectious. The population is well mixed, and the model assumes that latent and infectious times of the pathogen are exponentially distributed. In this letter, contact rate *β* is changing over time as it happens in COVID-19, which increase in early stage due to public unawareness of the disease, then decrease with government control policy measures. The contact rate follow a logistic function trend by estimating it day by day [9]:

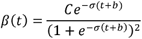

With *t* the number of days after first cases found in each region, for example in Lombardy January 31^st^ (the first found cases in Italy), *σ* a regularization parameter, and *b* the bias. A training procedure is performed on the observable data in order to find optimal (*C, σ, b*). We have considered as Exposed people, a number of twice infected people, after lockdown which is in line with the predictions and the observed values.

## Conclusions

We modeled spreading of Covid-19 using Chinese data and used the model to predict epidemic curve in each Italian region, allowing to gain better information on the new daily cases peaks with the forecasted curve. The forecast portion of the curve allows to have a better prediction of active cases with the SEIR model, by computing the position of the peaks of active cases for each Italian region. Training on Chinese data and use the knowledge to forecast Italian spreading of Covid-19 has resulted in a good fit, measured with the mean average precision between official Italian data and the forecast. SEIR model can gain advantage on modeling epidemic since the compartments are based on the complete curve dynamic (the portion of the real data and those forecasted). We showed the validity of the method since the predictive model learns from the dynamics of Covid-19 in China and exploits its learned knowledge to predict future daily cases in Italy. As shown in Figure 9 and in Table 2, the expected peak of SEIR model was confirmed at the end of March at national level. It is expected that Southern Italian Regions could reach the peak of total positive cases later, by April 11^th^, as estimated with dynamic SEIR model.

**Table 2.**
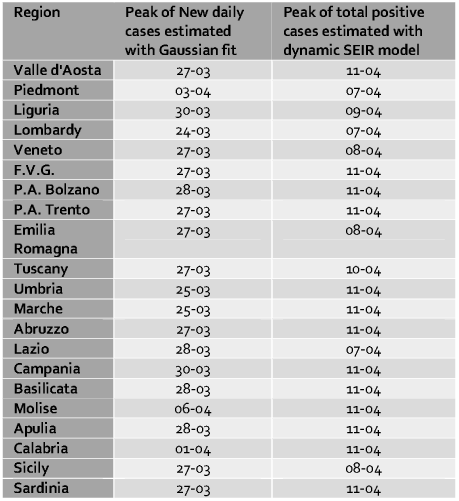
Estimation of peaks computed by forecasting the new daily cases with the modified autoencoder. The augmented time series is fit with a Gaussian function to find the date that occur for new daily cases, and the date where occur the total positive with the SEIR model. SEIR model here benefit from the forecasted data.

**Figure 8.**
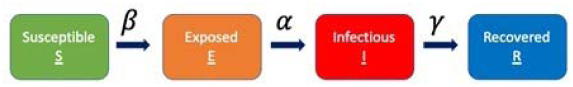
SEIR model with the four compartments and their relationships

**Figure 9.**
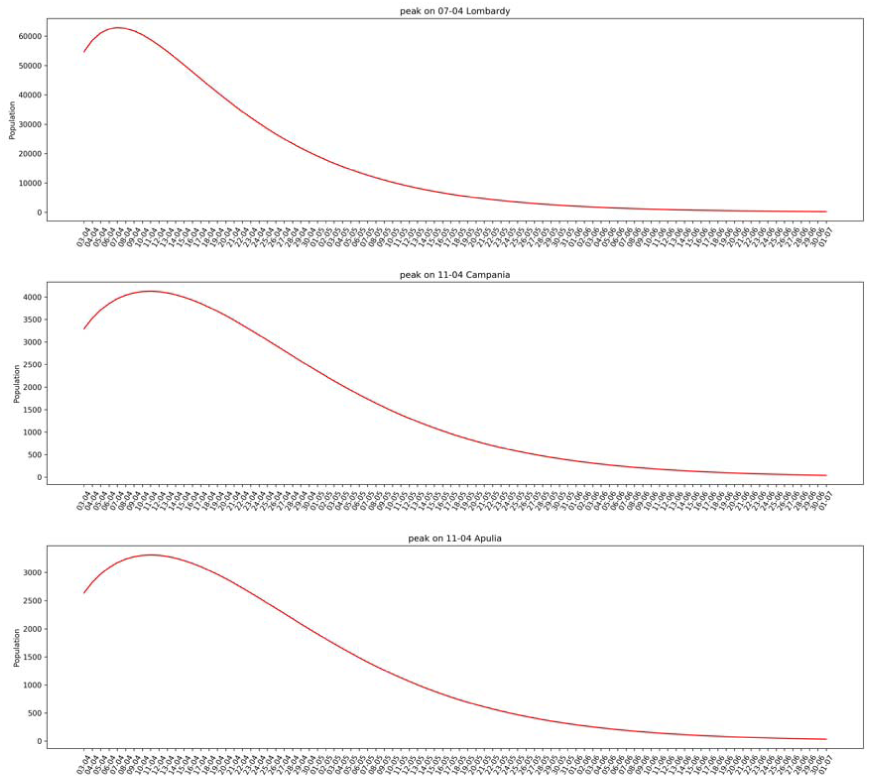
Prediction of the total positive for the two biggest Southern Italian Regions vs. Lombardy.

## Data Availability

Data are available from GitHub official website of the Department of "Protezione Civile", a structure of the Italian Prime Minister office

https://github.com/pcm-dpc/COVID-19

## Authors Contribution

AD, PP, AM conceived, wrote and revised the manuscript

